# Dysfunctional neuro-muscular mechanisms explain gradual gait changes in prodromal spastic paraplegia

**DOI:** 10.1101/2022.10.14.22281080

**Authors:** Christian Laßmann, Winfried Ilg, Tim W. Rattay, Ludger Schöls, Martin Giese, Daniel F.B. Haeufle

## Abstract

In Hereditary Spastic Paraplegia (HSP) type 4 (SPG4) a length-dependent axonal degeneration in the cortico-spinal tract leads to progressing symptoms of hyperreflexia, muscle weakness, and spasticity of lower extremities. Even before the manifestation of spastic gait, in the prodromal phase, axonal degeneration leads to subtle gait changes. These gait changes – depicted by digital gait recording – are related to disease severity in prodromal and early-to-moderate manifest SPG4 subjects. We hypothesize that dysfunctional neuro-muscular mechanisms such as hyperreflexia and muscle weakness explain these disease severity-related gait changes of prodromal and early-to-moderate manifest SPG4 subjects. We test our hypothesis in computer simulation with a neuro-muscular model of human walking. We introduce neuro-muscular dysfunction by gradually increasing sensory-motor reflex sensitivity based on increased velocity feedback and gradually increasing muscle weakness by reducing maximum isometric force. By increasing hyperreflexia of plantarflexor and dorsiflexor muscles, we found gradual muscular and kinematic changes in neuro-musculoskeletal simulations that are comparable to subtle gait changes found in prodromal SPG4 subjects. Predicting kinematic changes of prodromal and early-to-moderate manifest SPG4 subjects by gradual alterations of sensory-motor reflex sensitivity allows us to link gait as a directly accessible performance marker to emerging neuro-muscular changes for early therapeutic interventions.

## Introduction

In many neurodegenerative movement disorders like Parkinson’s disease, cerebellar ataxia, or hereditary spastic paraplegia (HSP), gait impairments are among the leading symptoms. They often appear as the first signs (***Globas et al., 2008***; ***Serrao et al., 2016***; ***Ilg et al., 2016***; ***Mirelman et al., 2016***) and are one of the most disabling features in the progression of these diseases. Recently, it has become possible to quantify specific subtle gait changes in early disease phases or even before the manifestation of clinical disease symptoms (***Ilg et al., 2016***; ***Mirelman et al., 2016***). The prodromal phase of movement disorders (***Rattay et al., 2022***) attracts increasing research interest, as it provides a promising window for early therapeutic intervention before substantially irreversible neurodegeneration has occurred.

We have recently shown for hereditary spastic paraplegia type 4 (SPG4) subjects — the most common autosomal dominant and pure motor form of HSP (***Rattay et al., 2022***; ***Hazan et al., 1999***) — that specific subtle changes in the kinematic gait pattern can be detected by quantitative movement analysis in the prodromal phase, before the manifestation of spastic gait (***Lassmann et al., 2022***). Changes that can be observed early are increased minimum plantarflexion or reduced foot range of motion (RoM) which gradually increase in early manifest stages (***Lassmann et al., 2022***), leading to gait patterns affecting the ankle, knee, and hip joints (***Martino et al., 2018, 2019***; ***Piccinini et al., 2011***). Especially the foot RoM and minimum plantarflexion show significant correlations to disease severity already in prodromal and early manifest stages (***Lassmann et al., 2022***).

On the neuro-muscular level, key pathologies observed in HSP patients are hyperreflexia, leg spasticity, and muscle weakness (***Fink, 2013***) caused by dying back axonopathy (***Rezende et al., 2015***; ***Lindig et al., 2015***). The origin of these pathologies is a length-dependent affection of the cortico-spinal tract (***Harding, 1983***; ***Fink, 2006***). Due to the length-dependency, early gait changes have been primarily observed in the ankle joint (***Lassmann et al., 2022***; ***Serrao et al., 2016***). Brisk patellar and Achilles reflexes can be observed in clinical examinations already in the prodromol phase (***Rattay et al., 2022***). In the manifest stage, additional spasticity and muscle weakness can be observed in static conditions as well as in gait ***Marsden et al. (2012***); ***Martino et al. (2019***); ***Rinaldi et al. (2017***). However, it is unknown to which part spastic hyperreflexia or muscle weakness contribute to the subtle gait changes observed in prodromal and early phases.

In order to understand the emerging gait abnormalities in early disease stages, it is crucial to investigate the development on the level of dysfunctional sensory-motor control mechanisms. Forward-dynamic computer simulation with neuro-musculoskeletal models allows for investigating the effect of isolated sensory-motor alterations (***De Groote and Falisse, 2021***). This method allows to reproduce healthy gait (***Geyer and Herr, 2010***; ***Song and Geyer, 2015***) and to study the contribution of individual sensory-motor reflexes to gait patterns (***van der Krogt et al., 2016***; ***Haeufle et al., 2018***; ***Schreff et al., 2022***; ***De Groote and Falisse, 2021***). The effect of neurodegenerative dying back axonopathy, as seen in HSP, on gait can be investigated by gradual manipulation of specific neuro-muscular mechanisms. Incremental bilateral plantarflexor weakness affecting gait was previously investigated by ***Waterval et al. (2021***). ***van der Krogt et al. (2010***) reproduced gait characteristics of children with cerebral palsy by introducing a velocity-dependent stretch reflex, increasing muscle activity for the fast stretch of muscle fibers. ***Jansen et al. (2014***) showed how hyperexcitability of muscle spindle reflex loops contribute to hemiparetic gait by investigating length- and velocity feedback. ***Bruel et al. (2022***) combine the effects of muscle weakness and hyperreflexia to explain the sensory-motor origin of the spastic heel- and toe-walking. In their study, they added muscle spindle-, length-, and force feedback to the two plantarflexor muscles, Soleus (SOL) and Gastrocnemius medialis (GAS), and introduced muscle weakness by reducing the maximum isometric muscle forces (***Bruel et al., 2022***).

In this study, we hypothesize that a gradual manipulation in sensory-motor reflex sensitivity and muscle weakness can explain the emergence of early gait changes in prodromal subjects towards early spastic gait in manifest SPG4 patients (see ***Figure 1*** for the study design). The gait of prodromal subjects and manifest SPG4 patients had an intact gait cycle structure consisting of heel strike, roll-over, push-off and swing phases (here called: heel strike walking). We base our approach on a previously published model predicting healthy human gait kinematics and dynamics (***Geyer and Herr, 2010***). In this model, we gradually manipulate hyperreflexia based on muscle spindle velocity feedback and muscle weakness to determine whether a singular neuro-muscular dysfunction or only their combination can explain the gradual kinematic changes observed in experimental data. We expect that developing gait changes over disease severity of prodromal subjects to the spastic gait of mild-to-moderate manifest patients can be predicted by altering plantarflexor and dorsiflexor muscle spindle reflex sensitivity and leg muscle weakness, caused by length-dependent axonal degeneration in SPG4.

**Figure 1.**
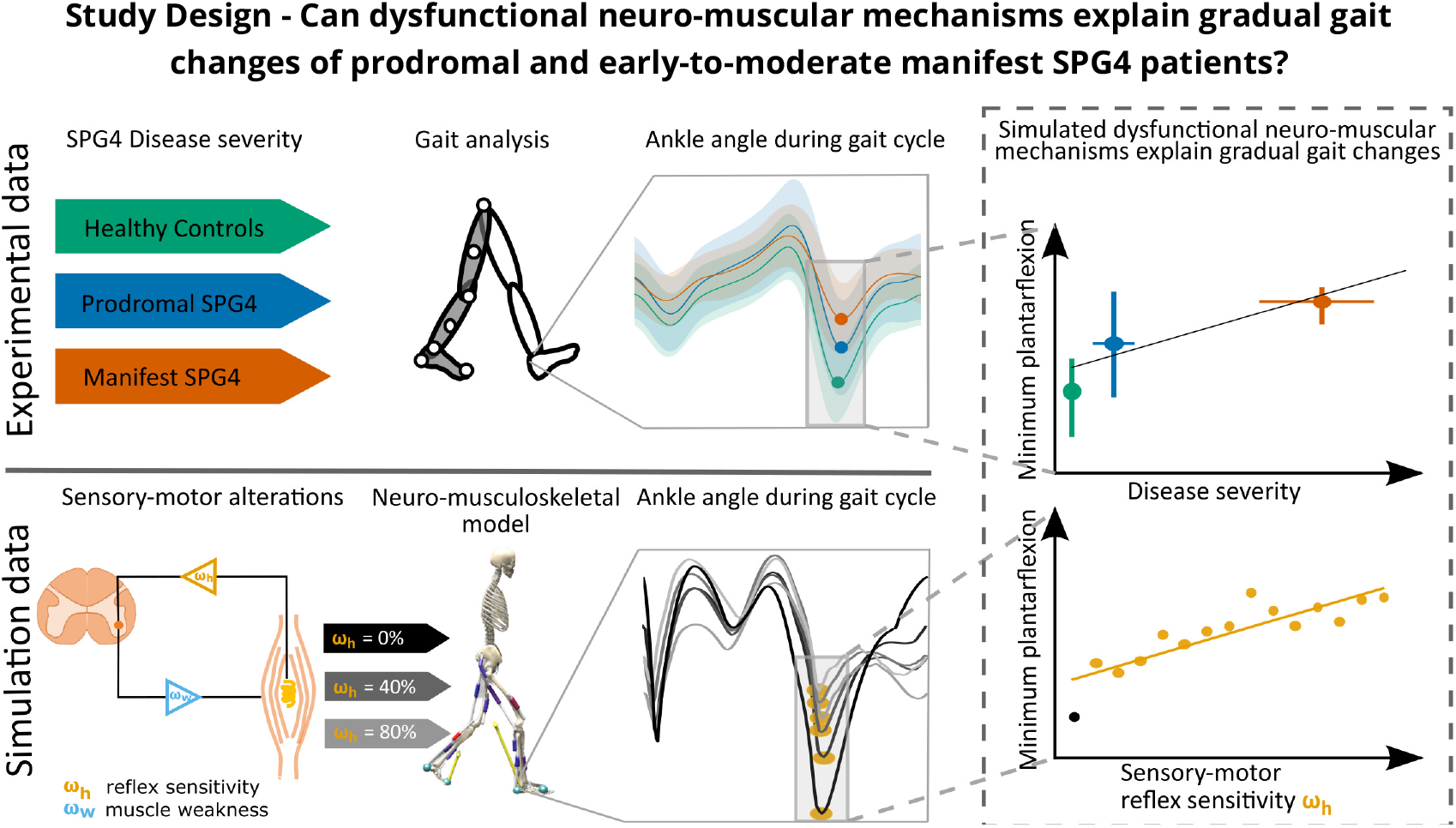
We first used data of an instrumented gait analysis to investigate gait changes of healthy controls (green), prodromal SPG4 (blue), and manifest SPG4 (red) patients. We identified characteristic changes for the three different groups, which were recently published (***Lassmann et al., 2022***). Secondly, we introduced and gradually manipulated neuro-muscular mechanisms, i.e. hyperreflexia (muscle spindle velocity feedback, orange), muscle weakness (reduced isometric force, light blue), and their combination in a neuro-musculoskeletal model and expected to predict relative gait changes, as in experimental data.

## Methods and Materials

### Experimental Data

We evaluate data from our previously published study (***Lassmann et al., 2022***), which included 17 manifest SPG4 patients, 30 prodromal SPG4, and 23 healthy control participants. The study was conducted according to the Helsinki Declaration and approved by the Institutional Review Board of the University of Tübingen (reference number: 266/2017BO2) for the preSPG4 Study. In addition, written informed consent was obtained from all study participants. Subjects were instructed to walk normally in a self-determined pace. All participants had an intact gait cycle structure consisting of heel strike, roll-over, push-off and swing phases (here called: heel strike walking). Participants underwent an instrumented gait analysis in a movement laboratory using an infraredcamera-based motion capture system (VICON FX with ten cameras). Gait cycles were recorded with 41 reflecting markers at a sampling rate of 120 Hz, and extracted by detection of the heel strike event. Trials were smoothed by a Savitzky-Golay polynomial filter and resampled equidistantly to 100 data points per gait cycle. For the analysis, we calculated stride length, gait speed and joint angles, to compare to simulated data.

### Computational model of human gait

We used a neuro-musculo-skeletal model of human gait, as it was used recently by ***Bruel et al. (2022***). The model is planar (sagittal plane) with seven segments (trunk-pelvis, bilateral thigh, lowerleg, and foot) and seven degrees of freedom (simplified from OpenSim gait2392 (***Delp et al., 2007***)). The planar model was used, since the most prominent differences between healthy controls, prodromal SPG4, and manifest SPG4 patients were found in the flexion and extension angles especially in the foot segment (***Lassmann et al., 2022***). We modeled seven Hill-type muscles (Millard-equilibrium muscle model (***Millard et al., 2013***)), namely Gluteus maximus (GLU), Iliopsoas (IL), Rectus femoris (RECT), Vastus intermedius (VAS), Gastrocnemius medialis (GAS), Soleus (SOL), and Tibilias anterior (TA) per leg. Muscle path, optimal fiber length, pennation angle, tendon slack length, and maximum isometric forces were set to the values in the Gait2392 model. Ground contact was modeled using two viscoelastic Hunt-Crossley contact spheres on each foot, serving as heel and toe contacts.

The neuronal control model calculated muscle stimulation signals *U* for each of the fourteen muscles according to a gait state-dependent reflex-based controller based on ***Geyer and Herr*** (***2010***). The controller considered muscle force and length feedback, vestibular feedback, and constant signals to generate muscle excitation. Reflex gains could differ between five gait phases (early stance, late stance, lift-off, swing, and landing), as proposed in previous studies (***Song and Geyer, 2015***; ***Ong et al., 2019***; ***Waterval et al., 2021***).

### Simulation study design: a model of spastic hyperreflexia and muscle weakness

This study systematically introduced sensory-motor alterations to model healthy, prodromal and early-to-moderate manifest gait in SPG4. The study design has two axes. On the first axis, we investigated three different control scenarios: spastic hyperreflexia, muscle weakness, and a combination of both. On the second axis, we investigated the magnitude of the respective sensory-motor alterations.

#### First axis

To model spastic **hyperreflexia**, we introduced a gain parameter *ω*_*h*_ ∈ [0%…100%]. *ω*_*h*_ is multiplied by the equation calculating the muscle spindle velocity feedback:

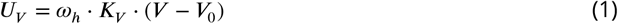

with *V* and *V*_0_ being the normalized CE velocity ((*L*/*L*_*opt*_)/ *s*) and the respective constant reference velocity. *K*_*V*_ = 0.12 s is the velocity feedback gain which did not lead to a walking gait in our optimization approach in an exploratory examination. *ω*_*h*_ = 0% results in a deactivated velocity reflex and *ω*_*h*_ = 100% results in the maximally investigated velocity reflex sensitivity (hyperreflexia). *ω*_*h*_ was added to the ankle plantarflexors GAS and SOL, and ankle dorsiflexor TA during the stance and swing phase.

To model **muscle weakness**, we introduced a gain parameter *ω*_*w*_ ∈ [0%…100%] which directly reduces the maximum isometric muscle force (*F*_max_) of the leg muscles. *ω*_*w*_ = 0% represents a model with all muscles at full strength, while *ω*_*w*_ = 100% represents a reduction of the isometric force to levels reported by ***Marsden et al. (2012***), namely 42% of dorsiflexors, 58% of plantarflexors, 62% knee extensors, 65% knee flexors, 89% hip extensors, and 70% hip flexors.

To model the third scenario, we combined both approaches to investigate the interplay of both symptoms. For this, we introduced the parameter *ω*_*hw*_, which sets both, velocity feedback gain *ω*_*h*_ = *ω*_*hw*_ and muscle weakness *ω*_*w*_ = *ω*_*hw*_ simultaneously.

#### Second axis

To investigate gradual sensory-motor alterations, the magnitude of the gains was increased in 15 steps: *ωh, w, hw* = [0%, 6.67%, 13.34%, 20%, …, 100%]. Low *ω*-values mean minimal sensory-motor alterations, i.e., low hyperreflexia and muscle weakness, while *ω*-values of 100% represent the highest alterations investigated in this study. See Supplementary Figure 1 for details on the gradual change of velocity feedback gain and muscle weakness and their combination.

### Optimization of controller parameters

For each of the scenarios described above, all other controller parameters were optimized. These are the feedback gains of the other reflexes (length, force, and vestibular) within each state, transition thresholds between the phases, and the initial joint angles. We used the open-source software SCONE with Hyfydy for the optimization, a dedicated software to run and optimize predictive neuromuscular simulations ***Geijtenbeek*** (***2019***, 2021). The cost function for the optimization (see ***Equation 2***) considered a minimum gait speed ***Equation 3***, an effort measure from ***Wang et al. (2012***) minimizing metabolic energy expenditure of muscles (*J*_effort_), a joint measure penalizing hyperextension and -flexion of the ankle (***Equation 4***) and knee (***Equation 5***) joints, and ground reaction force measure penalizing high forces during gait (*J*_grf_):

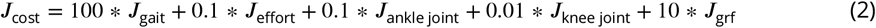

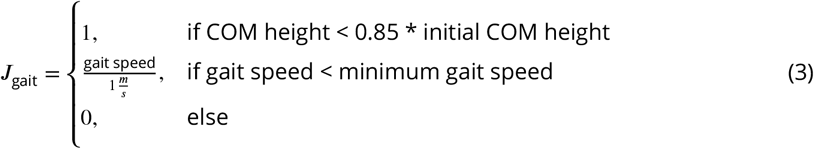

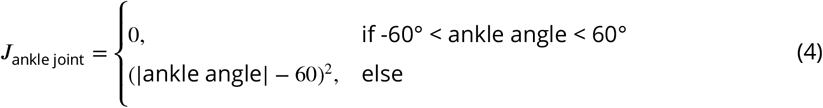

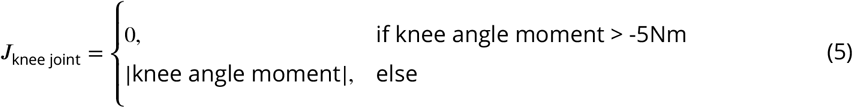

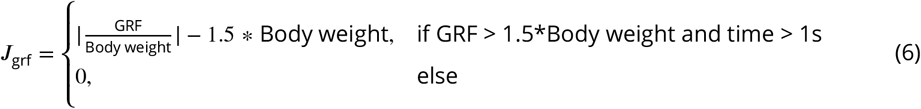

SCONE uses the Covariance Matrix Adaptation Evolutionary Strategy from ***Igel et al. (2006***). The optimization was stopped when the average reduction of the cost function score was less than 0.0001% compared to the previous iteration. We simulated gait for 30 seconds, always starting from the same initial parameters. We only considered simulations with stable walking until the simulation ends. For analysis we excluded the first gait cycle, resampled to 100 data points per gait cycle, and averaged over all gait cycles.

### Data evaluation

We compared the simulation output to the experimental data for specific relevant gait features identified in our previous study ***Lassmann et al. (2022***). As gradually altering features in prodromal and manifest SPG4, we identified the minimum plantarflexion, the foot range of motion (RoM), and the maximum ground clearance of the heel. For manifest SPG4 the knee angle at heel strike increased and the maximum heel angle and knee RoM reduced significantly. Furthermore, gait speed and stride length were reduced over disease progression for manifest SPG4 patients ***Lassmann et al. (2022***). Hip, knee, and ankle joint angle kinematics during the gait cycle were compared between healthy controls, prodromal SPG4 subjects, and manifest SPG4 patients. We compared the simulation results with nine key features of the experimental data: (1) ankle RoM, (2) minimum plantarflexion (swing phase), (3) ankle angle at heel strike, (4) ankle angle at maximum heel ground clearance, (5) knee RoM, (6) maximum knee angle, (7) knee angle at heel strike, (8) gait speed, and (9) stride length. Peak and average muscle activation for SOL, GAS, and TA was calculated for each gait phase (early stance, late stance, lift-off, swing, and landing). SOL and TA co-activation values were calculated with average muscle activation values for each of the five gait phases:

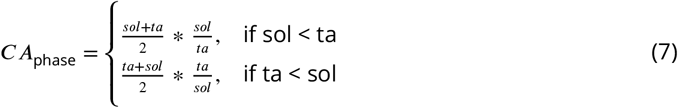

where sol and ta represent the mean muscle activation for a certain gait phase. For statistical comparison Kruskal-Wallis test and post hoc Dunn’s test for multiple group comparisons were used. We report statistical significance as *: p<0.05, **: p<0.0056 (Bonferroni corrected with 9 feature comparisons), and ***: p<0.001.

We used the SPRS score (***Schüle et al., 2006***) to categorize subjects into clinical disease severity and find possible explanations by increasing velocity feedback gains in the simulations.

Spearman’s *rho* was used to identify significant correlations of increased muscle spindle velocity feedback and increased muscle weakness for the nine gait features and optimization parameters, e.g., force feedback gains of individual muscles and metabolic energy expenditure (***Wang et al., 2012***).

## Results

### Experimental gait data

As recently published, instrumental gait analysis revealed significant group differences between healthy controls (HC), prodromal SPG4 subjects, and manifest SPG4 patients (***Lassmann et al., 2022***). All participants performed a self-determined heel strike walking. For this study, we extracted joint angle kinematics and other gait parameters, as described in detail by ***Lassmann et al. (2022***). Several gait parameters showed significant differences between healthy controls and prodromal SPG4 with increasing effects in manifest SPG4 patients. Minimum plantarflexion (p=0.029*), and ankle angle at maximum heel ground clearance (p=0.029*) were significantly increased for prodromal SPG4 and manifest (p<0.001***) in comparison to healthy controls. Pearson’s rho showed a gradual increase of these features with disease severity (rho=0.48, p<0.001***; rho=0.5, p<0.001***, respectively). For manifest SPG4 patients, the ankle and knee RoM (p<0.001***) and maximum knee angle (p=0.013*) were significantly reduced, and the knee angle at heel strike was increased (p<0.001***). The gait speed and stride length were decreased for manifest SPG4, but not for prodromal SPG4 subjects. ***Table 1*** shows mean values and standard deviation for all nine analyzed features of the three groups.

**Table 1.**
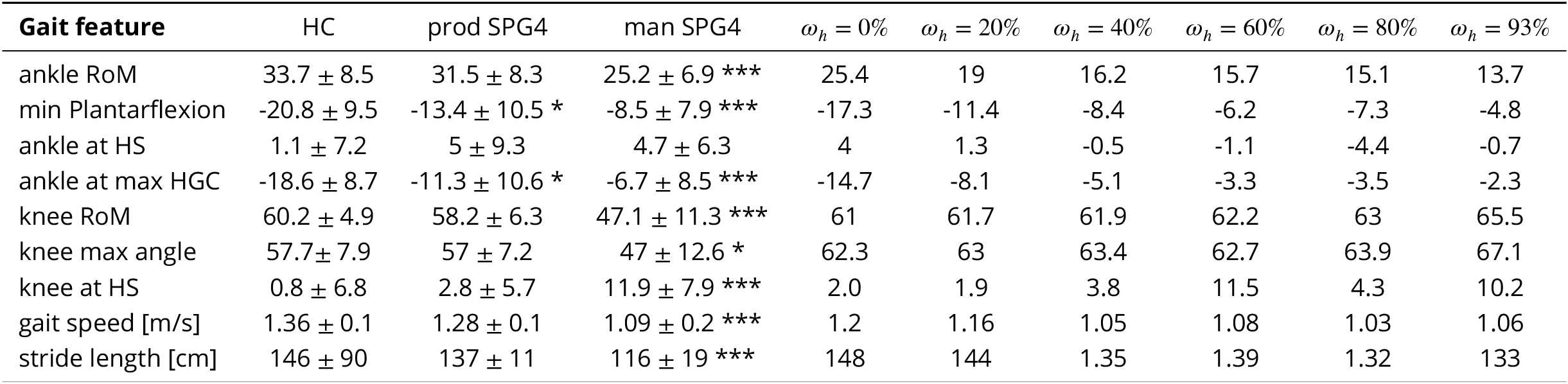
Mean results for gait features with standard deviation (STD) for experimental and simulated data. RoM ≡ Range of Motion, PF ≡ Plantarflexion, HS ≡ Heel strike, HGC ≡ Heel ground clearance. Asterisks indicate significance: * if *p* < 0.05 and ** if *p* < 0.001 in comparison to HC. Different levels of the velocity feedback gain scenario (*ω*_*h*_) are given for comparison.

Kinematics of the ankle, knee, and hip joint during the gait cycle showed differences between HC (green), prodromal SPG4 (blue), and manifest SPG4 (red in ***Figure 2a-c***). The most prominent differences occurred during the swing phase, e.g., the increasing minimum plantarflexion angle from healthy controls to prodromal subjects and manifest SPG4 patients (***Figure 2a*** at around 70% of the gait cycle), indicating a progression with disease severity. Furthermore, the increased knee angle at heel strike in the manifest group is visible (***Figure 2b***, the beginning of the gait cycle).

**Figure 2.**
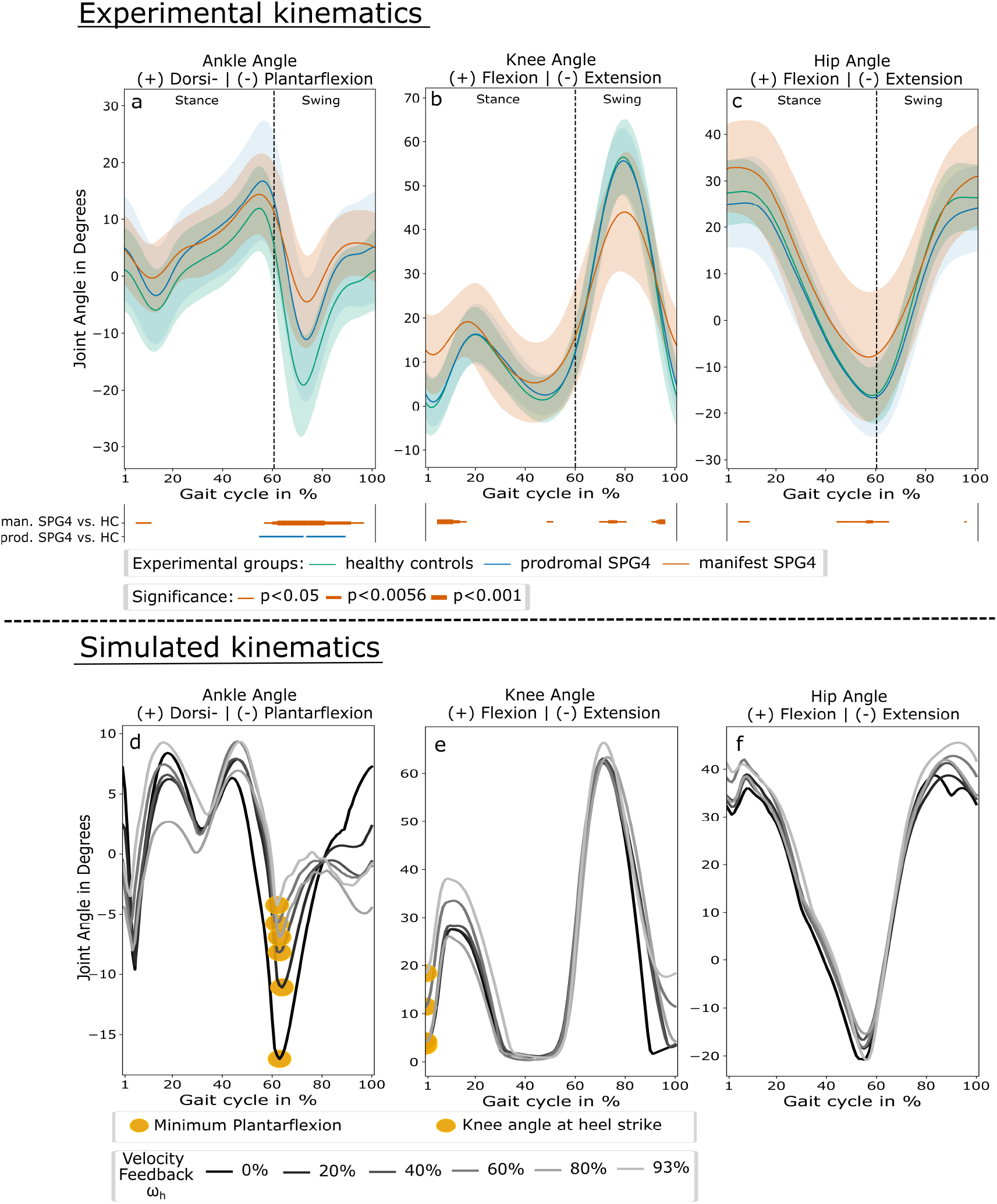
a-c: mean flexion and extension angles of ankle, knee, and hip joints over the gait cycle in percent for healthy controls (green), prodromal SPG4 (blue), and manifest SPG4 (red), with their standard deviation. Significant periods are indicated as lines above the trajectory plots indicating different levels of significance (thin line: p<0.05, intermediate line: p<0.0056, and bold line: p<0.001. Differences between prodromal SPG4 vs. HC and manifest SPG4 vs. HC are shown as blue and red lines, respectively. d-f: flexion and extension angles of ankle, knee, and hip joints over the gait cycle in percent for different levels of velocity feedback gains (color coded from black: *ω*_*h*_ = 0%, light grey: *ω*_*h*_ = 93%) of plantarflexor and dorsiflexor muscles. The extracted features are highlighted yellow, namely the minimum plantarflexion (d) and knee angle at heel strike (e).

### Neuro-musculoskeletal gait model

#### Simulated healthy walking pattern

The simulation of the not adapted ***Geyer and Herr*** (***2010***) controller can reproduce healthy gait. ***Figure 2d-f*** in black (*ω* = 0%) and ***Table 1*** show the results for the model with optimized controller parameters (optimized in Scone). We found reduced maximum ankle dorsiflexion and a more extended swing phase compared to our experimental data.

#### Effect of increasing velocity feedback gain

With increasing levels of velocity feedback gain *K*_*V*_ (*ω*_*h*_) to plantarflexor and dorsiflexor muscles during the stance and swing phase, several kinematic changes occurred within heel strike walking.

#### Ankle

The minimum plantarflexion angle reduced from -17.3° (*ω*_*h*_ = 0%) to -11.4° at *ω*_*h*_ = 20% and further to -8.35° (*ω*_*h*_ = 40%) and -4.8° (*ω*_*h*_ = 93%) (see ***Figure 2d***). This resulted in a strong correlation between increasing velocity feedback gains and minimum plantarflexion (rho=0.9, p<0.001***, compare ***Figure 3a***). In addition, also the ankle angle at heel strike was gradually increased (rho=- 0.87, p<0.001***) and gait speed was reduced (rho=-0.53, p=0.04*). **Knee:** At heel strike, the knee angle was gradually increased from *ω*_*h*_ ≥ 53% to *ω*_*h*_ = 93% (rho=0.88, p<0.001***, compare ***Figure 2e*** and ***Figure 3b***). For comparison with experimental data, the results of different iterations of increasing velocity feedback gain are shown in ***Table 1*** and all results with correlations in Supplementary Table 1.

**Figure 3.**
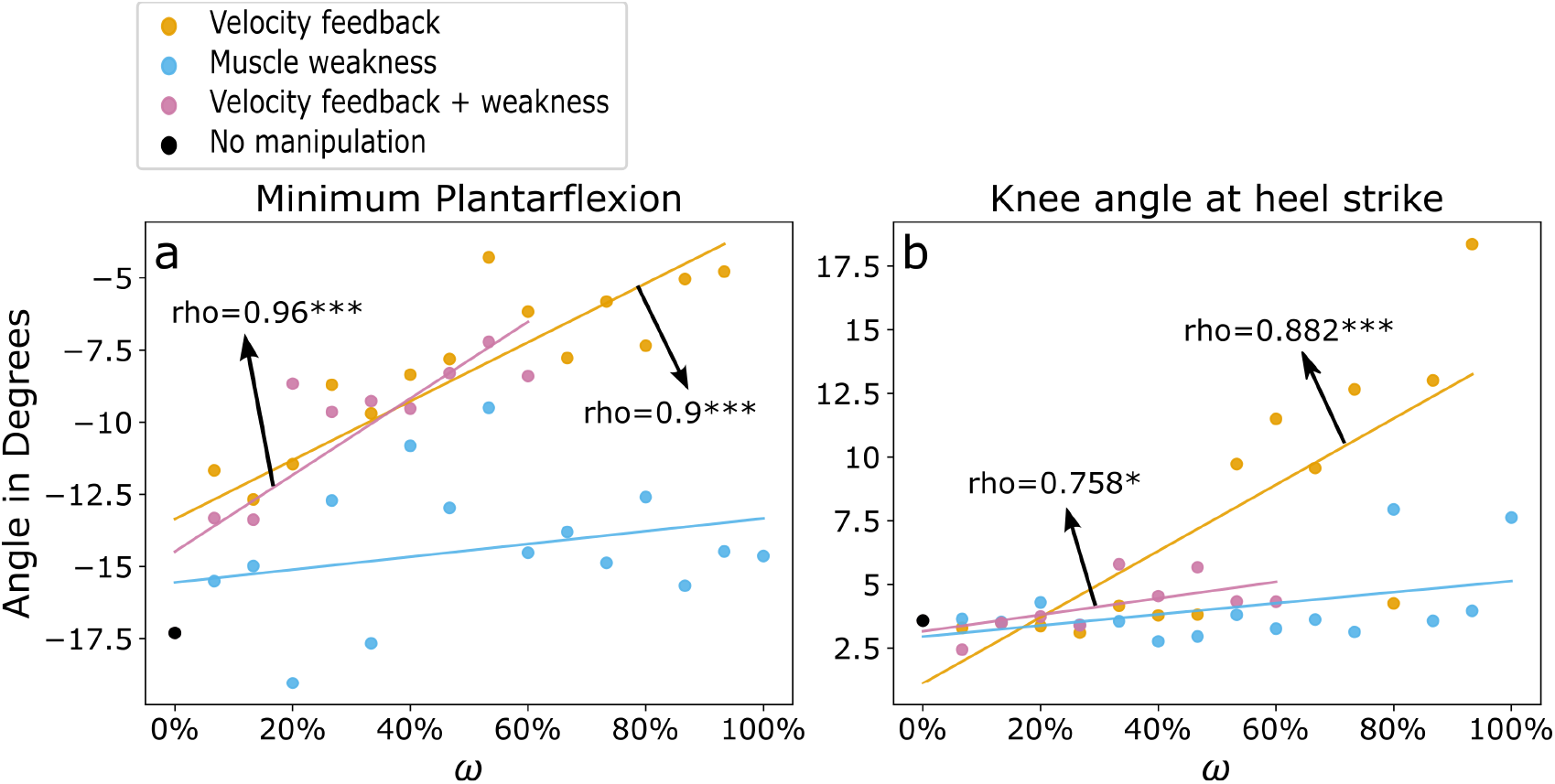
Increasing levels of velocity feedback gain (orange), muscle weakness (light blue), and velocity feedback gain + muscle weakness (purple) with simulation iteration *ω* = 0% to *ω* = 100% and linear fits. a) minimum plantarflexion and b) knee angle at heel strike are shown with significant pearson correlation coefficients. Asterisks indicate significant levels of: *: p<0.05, **: p<0.0056, and ***: p<0.001. For *ω*_*h*_ = 100% and *ω*_*hw*_ > 60% optimization led to no stable walking simulations.

SOL average activation was increased during the early stance phase (rho=0.95, p<0.001) and reduced during lift-off (rho=-0.75, p=0.0012) Supplementary Figure 2a. During landing, there was a greater SOL activation (rho=0.84, p<0.001). For GAS, the average activation during the early stance phase was increased with increasing *ω*_*h*_, showing a prolonged GAS activation over the stance phase; however, with a shortened peak muscle activation period Supplementary Figure 2b. During the landing phase, the GAS average activation increased with higher velocity feedback gain (rho=0.97, p<0.001). Tibialis anterior (TA) peak activation increased at early stance (rho=0.87, p<0.001) Supplementary Figure 2c. During swing and landing, TA activity increased with *ω*_*h*_ (rho=0.68, p=0.0057; rho=0.94, p<0.001, respectively) Supplementary Figure 2c. SOL-TA co-activation increased during early stance (rho=0.87, p<0.001), swing (rho=0.68, p=0.0057), and landing (rho=0.93, p<0.001), and decreased during lift-off (rho=-0.63, p=0.013) with increasing *ω*_*h*_.

All iterations with increasing muscle spindle velocity feedback gain, except for *ω*_*h*_ = 100%, could be optimized to a stable walking simulation.

#### Effect of increasing muscle weakness

The gradual increase of muscle weakness *F*_max_ (*ω*_*w*_) as reported in ***Marsden et al. (2012***) resulted in an increased ankle angle at heel strike (rho=0.8, p<0.001***, compare ***Figure 3a***). The maximum knee angle differed between simulation scenarios in a range of 52° (*ω*_*w*_ = 20%) and 75° (*ω*_*w*_ = 66.7%), with no significant correlation over increased muscle weakness. Other investigated features did not show a specific pattern with increasing muscle weakness. All simulations with increasing muscle weakness (*ω*_*w*_ = 0%…100%) could be optimized to a stable heel strike walking simulation. For all simulation results, see Supplementary Table 2 and Supplementary Figure 3.

#### Combined velocity feedback gain and muscle weakness

The combination of a gradual increase of velocity feedback gain and muscle weakness (*ω*_*hw*_) resulted in patterns similar to the velocity feedback gain scenario. During the swing phase, the minimum plantarflexion was reduced for higher *ω*_*hw*_ (rho=0.96,p<0.001***, see ***Figure 3a***). The ankle angle reduced at heel strike (rho=-0.96,p<0.001***) and increased at maximum heel ground clearance (rho=0.98,p<0.001***). The knee angle at heel strike increased with *ω*_*hw*_ (rho=0.76,p=0.011*, see ***Figure 3b***). Gait speed and stride length were reduced to comparable levels as in the velocity feedback gain scenario, however, with no significant correlation to increased *ω*_*hw*_ (see Supplementary Table 3 and Supplementary Figure 4). The optimizer failed to produce stable heel strike walking with *ω*_*hw*_ ≥ 60%, showing a reinforced effect by combining the gradually increased velocity feedback gain and muscle weakness. At *ω*_*hw*_ = 73% the optimization dismissed the heel strike walking but produced a stable toe-walking pattern with initial ball contact, increased hip flexion angle and a time offset at maximum knee flexion angle (compare Supplementary Figure 5).

#### Optimized control parameters and cost terms

For each specified velocity feedback gain and/or muscle weakness parameters (*ω*_*h*_, *ω*_*w*_, or *ω*_*hw*_), we optimized all other controller parameters to find a suitable gait minimizing our locomotion cost function (***Equation 2***). This re-optimization resulted in changes in the cost terms and the controller parameters and reflected the possibility of the rest of the nervous system adapting to specific sensory-motor changes. For increasing velocity feedback gain and combined velocity feedback gain and muscle weakness, the cost term *J*_effort_ metabolic energy expenditure (***Wang et al., 2012***) increased with increasing *ω* (*ω*_*h*_ : *rho* = 0.79, *p* < 0.001***; *ω*_*hw*_ : *rho* = 0.66, *p* = 0.038*, respectively). The controller parameter *length feedback gain* of TA (optimized over the whole gait cycle) increased with higher velocity feedback gains (*rho* = 0.72, *p* = 0.002**). The *force feedback gains* of SOL and GAS during lift-off and swing phases decreased with higher velocity feedback gains (*rho* = −0.94, *p* < 0.001***, for both) and combined velocity feedback and muscle weakness (*rho*_SOL_ = −0.98, *p*_SOL_ < 0.001***; *rho*_GAS_ = −0.99, *p*_GAS_ < 0.001***). For the combined controller of muscle weakness and velocity feedback gain, the offset of TA muscle spindle length feedback (*L*_0_) was optimized to an increased value of 1.07 (as a fraction of the optimal TA muscle fiber length of 9.8cm) for the toe-gait scenario (*ω*_*hw*_ = 73%), in comparison: for all other combined controller scenarios (*L*_0_(*ω*_*hw*_ ∈ [6.67%…60%]) = 0.65 ± 0.003). This offset leads to a reduced TA activation during stance, lift-off, and landing Supplementary Figure 6. For more details on the optimized parameters, see Supplementary Table 4.

## Discussion

We hypothesized, that the subtle gait changes in heel strike walking observed in prodromal SPG4 subjects could be explained by gradual changes in neuro-muscular feedback mechanisms. To investigate this, we implemented gradually increased sensitivity of sensory-motor reflex in a neuromusculoskeletal forward simulation of heel strike walking (***Geyer and Herr, 2010***). Increasing levels of velocity feedback gain in plantarflexor and dorsiflexor muscles resulted in kinematic and muscular changes comparable to those observed in prodromal subjects and early-to-moderate manifest SPG4 patients.

### Increasing hyperreflexia explains the development of early gait changes in SPG4

On the kinematic level, the earliest gait changes in prodromal SPG4 subjects occur in the foot segment and ankle joint (***Lassmann et al., 2022***). Increasing muscle spindle velocity feedback (*ω*_*h*_) in the simulation caused several gait changes that are in line with kinematic changes of heel strike walking in prodromal subjects and early-to-moderate manifest SPG4 patients.

In the simulation, the minimum plantarflexion increased gradually with *ω*_*h*_ (rho=0.9, p<0.001) to comparable levels as it increased over disease severity, measured by the SPRS score (***Schüle et al., 2006***), in the experimental data of prodromal and early-to-moderate manifest SPG4 subjects (rho=0.49, p<0.001). With *ω*_*h*_ ≥ 53% the minimum plantarflexion saturates, as it has been shown in ***Lassmann et al. (2022***) for early-to-moderate manifest SPG4 patients.

The ankle RoM was identified as key feature of kinematic changes in prodromal and manifest SPG4 subjects (***Lassmann et al., 2022***) and used to cluster manifest HSP patients into severity-related groups (***Serrao et al., 2016***). In the simulation, the ankle RoM reduced gradually with increasing *ω*_*h*_ (rho=-0.99, p<0.001), as in the experimental data with disease severity (rho=-0.5, p<0.001). However, the absolute values did not fit the experimental data due to reduced maximum dorsiflexion in all simulations.

Comparable to the experimental data with disease severity (rho=0.48, p<0.001), the knee angle at heel strike was gradually increased with greater velocity feedback gain (rho=0.88, p<0.001). For low velocity feedback gains (*ω*_*h*_ < 53%), the knee angle at heel strike remained on a constant level comparable to healthy controls and prodromal SPG4 subjects. With greater velocity feedback gains, the knee angle at heel strike increased, matching the kinematic changes in manifest SPG4 patients.

Currently, there is no measurement or biomarker linking our velocity feedback gain parameter *ω*_*h*_ to disease severity. However, when plotting kinematic features like minimum plantarflexion and knee angle at heel strike of experimental data over SPRS score, which indicates disease severity (***Schüle et al., 2006***), and of simulated data over sensory-motor reflex sensitivity *ω*_*h*_, the plot suggests reproducing the gradually changing gait features of prodromal and early-to-moderate manifest SPG4 subjects with disease severity (see ***Figure 4***). These findings allow us to conclude that velocity-dependent hyperreflexia can explain the development of earliest gait changes in prodromal subjects and early spastic gait in patients with hereditary spastic paraplegia type 4 and shows the importance of gait as directly accessible performance marker for early therapeutic interventions.

**Figure 4.**
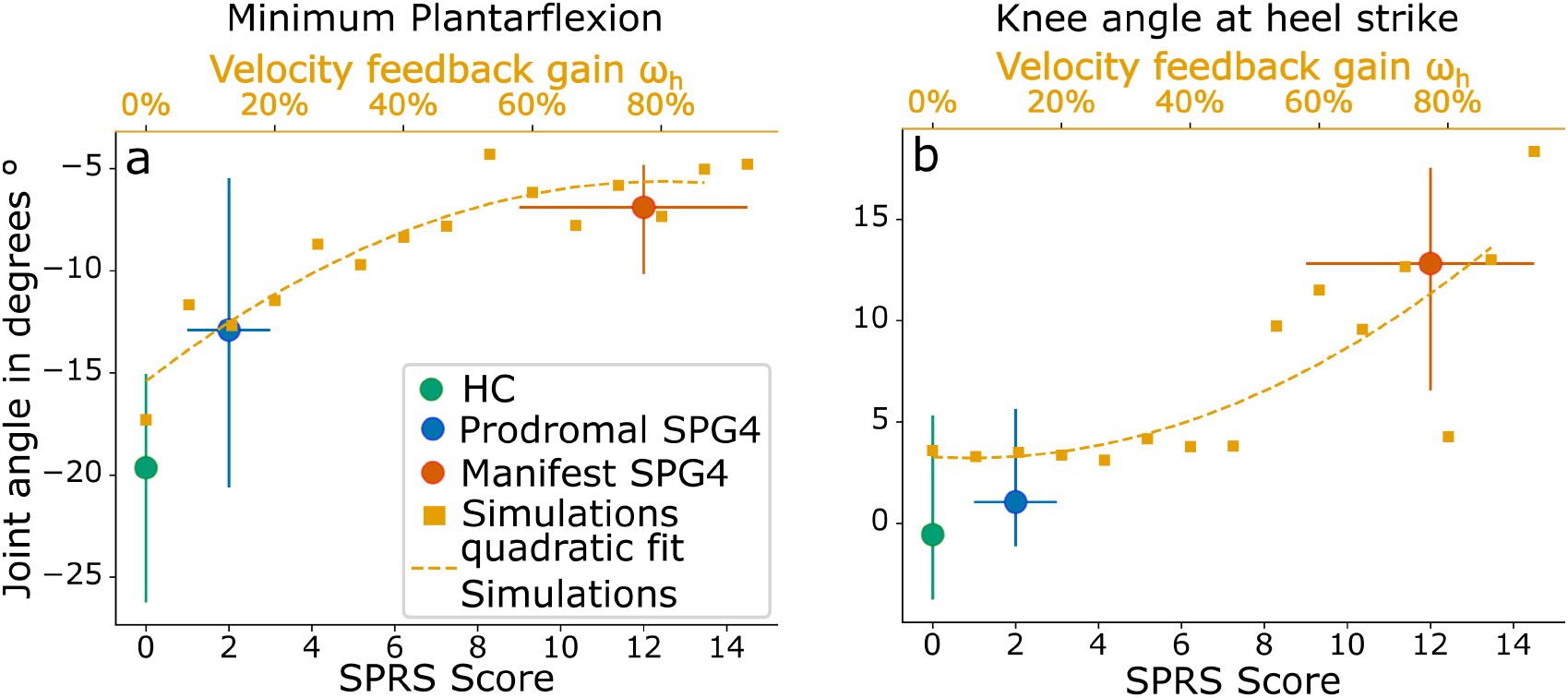
a: Minimum plantarflexion and b: knee angle at heel strike of experimental data and simulations over disease severity (SPRS score) and velocity feedback gain *ω*_*h*_. The three experimental groups are color-coded with healthy controls (green), prodromal SPG4 (blue), and manifest SPG4 (red). Shown are averaged values for SPRS scores as blue and red circles. Error bars are showing distributions of all groups with their mean SPRS score (position on lower x-axis) and standard deviation of SPRS score indicated by horizontal error bars. Orange squares are showing simulation data at different gains of velocity feedback (*ω*_*h*_, upper x-axis). Quadratic fit for simulations with increasing velocity feedback gain is shown in the respective color.

### Increasing hyperreflexia predicts changes in muscular coordination

The increasing velocity feedback gain *ω*_*h*_ has consequences beyond the kinematic changes. Optimizing all other neuronal control parameters for any given *ω*_*h*_, increased SOL and TA activity during the early stance and swing phase, with a higher level of co-activation during early stance and swing phase. ***Rinaldi et al. (2017***) reported a similarly increased co-activation of antagonist ankle muscles (SOL-TA) during the stance and swing phase in manifest HSP patients. ***Martino et al. (2019***) found a prolonged activation of ankle plantarflexor muscles in manifest HSP, which could be replicated in our simulated GAS activation, however, with a shorter peak period.

Our simulations’ metabolic energy expenditure (***Wang et al., 2012***) was positively correlated with increasing velocity feedback gains. This result is in line with ***Rinaldi et al. (2017***), who report an increase in energetic consumption in manifest HSP patients.

These findings indicate that the increased velocity feedback gain, a model representation of hyperreflexia in the ankle joint muscles, predicts not only kinematic but also muscular and energetic trends observed in prodromal and early-to-moderate manifest SPG4 patients.

### Severely spastic gait in manifest SPG4

In contrast to other simulation studies that focus on severe manifest spastic gait with altered gait patterns, we investigated the prodromal and early phases of spastic gait with an intact gait cycle structure consisting of heel strike, roll-over, push-off, and swing phases (here called: heel strike walking).

We did not find the kinematic changes occurring in prodromal and early-to-moderate manifest SPG4 subjects for increasing muscle weakness. However, the combined effects of hyperreflexia and muscle weakness *ω*_*hw*_ show the importance of muscle weakness in manifest hereditary spastic paraplegia. By simultaneously increasing both velocity feedback gain and muscle weakness, we found a toe-gait pattern Supplementary Figure 5, which is characteristic of later manifest stages of hereditary spastic paraplegia. Our results suggest a decrease of TA activation in the toe-gait scenario, resulting in a decrease of TA-SOL reciprocal inhibition. In combination with the increased plantarflexor velocity feedback gain, this leads to an over-activity of plantarflexor muscles during the stance and swing phase.

Other simulation studies previously investigated severe manifest gait in different movement disorders by introducing hyperreflexia and muscle weakness. ***Waterval et al. (2021***) simulated bilateral plantarflexor weakness by incrementally introducing GAS and SOL muscle weakness. They report that gait altered meaningfully when maximum isometric muscle force was reduced to less than 40%. In our study, we reduced muscle force to levels found by ***Marsden et al. (2012***), with a minimum muscle force of 42% occurring in dorsiflexors. We found no exclusive effect of the investigated muscle weakness on pathological gait in SPG4 patients, which might be explained by the still remaining isometric force of more than 40%. ***Bruel et al. (2022***) showed that increased velocity- and force-related sensory-motor reflexes of GAS and SOL lead to pathological toe-walking patterns, which can be seen in later stages of manifest spastic patients. Furthermore, ***Jansen et al. (2014***) used hyper-excitability of muscle spindle length- and velocity reflex loops to simulate hemiparetic gait in a neuro-musculoskeletal model. They found that both feedback mechanisms introduced to SOL, GAS, Vastus (VAS), and Rectus femoris (RECT), can lead to specific gait impairments, such as reduction of ankle dorsiflexion and decreased knee flexion during stance.

### Study limitations

In the combined sensory-motor reflex scenario of increased velocity feedback gain and muscle weakness, we assumed a simultaneous linear development of both factors from 0% to 100%. The experimental results of ***Rattay et al. (2022***) suggest that lower leg spasticity and muscle weakness emerge contiguously, but later than hyperreflexia, which was found in almost all prodromal SPG4 subjects (***Rattay et al., 2022***; ***Lassmann et al., 2022***). For higher *ω*_*hw*_ in the combined scenario, several optimizations did not find a stable walking gait. Further investigations in the longitudinal development of muscle weakness and hyperexcitability of muscle spindle reflex loops in SPG4 patients are necessary to understand the interplay of these symptoms.

The cost function for the parameter optimization determines the resulting gait pattern. For our simulations, we used a combined cost function that penalized excessive ground reaction forces, as suggested by ***Veerkamp et al. (2021***). Furthermore, the minimum gait speed was set to 1 m/s, which is the average gait speed of our early-to-moderate SPG4 group (***Lassmann et al., 2022***). Hyperextension and -flexion of ankle and knee joints were penalized to ensure normal gait patterns. We introduced this cost function, since we were interested in the subtle gait changes of prodromal and early-to-moderate SPG4 subjects, who still perform a heel strike walking pattern. To simulate more severe stages of SPG4, a different cost function may be needed, to allow a less constrained gait pattern (***Bruel et al., 2022***).

The length-dependent axonal degeneration in the cortico-spinal tract of SPG4 patients (***Fink, 2006***) suggests that spinal reflex changes may emerge first for distal reflex loops. For this reason, we studied gradual velocity feedback gains only at the most distal muscles (GAS, SOL, and TA). Also ***Martino et al. (2019***) found altered muscle activation in the most distal muscles. For muscle weakness, we considered an affection of all simulated muscles, as reported in ***Marsden et al. (2012***). Altering the sensory-motor reflex sensitivity in more proximal muscles may increase the simulation prediction accuracy of kinematic changes also in the other joints – at the cost of interpretation complexity. Nevertheless, it is crucial to investigate further the impact of muscle activation and hyperexcitability of the knee and hip muscle reflex loops, e.g., as ***Di Russo et al. (2021***) did to investigate the effect of different sensory-motor reflex sensitivities on gait speed and stride length.

The model we used is limited to simulating walking in the sagittal plane (two-dimensional). In severe manifest SPG4 patients, hip adductor spasticity is a common symptom (***Van Lith et al., 2019***) and leads to instability. Simulating the 3D gait pattern of SPG4 patients would be needed to include a more detailed symptomatic pattern of muscle spasticity and weakness.

### Conclusion and outlook

Very early kinematic changes in the gait pattern present a directly accessible performance measure for prodromal and manifest SPG4 subjects ***Lassmann et al. (2022***). We here identified sensorymotor reflex sensitivity changes as a possible explanation for these subtle kinematic changes. In our model, the gradual increase of reflex sensitivity can explain the gradual change in heel strike walking observed with increasing disease severity. On the other hand, muscle weakness could be compensated by other adapting spinal reflexes and did not lead to the observed kinematic changes. From this, we speculate that early pharmacological interventions to reduce spasticity (e.g., by baclofen) might reduce subtle gait changes by reducing the sensory-motor reflex sensitivity. However, the side-effects of increased muscle weakness may be compensated intraindividual through adapting spinal reflexes. This thought experiment indicates that pharmacological reduction of spasticity in early SPG4 patients could delay the onset of manifest spastic gait. In the currently running longitudinal experimental study (***Rattay et al., 2022***; ***Lassmann et al., 2022***), we will further investigate individual kinematic changes over time and simulate the development of sensory-motor reflex alterations to link gait changes to neuro-muscular mechanisms for future therapeutic interventions. Further studies are needed to objectively measure altered sensory-motor reflex loops and axonal damage in prodromal and early-to-moderate SPG4 subjects, e.g., a dynamometer-based H-reflex measure and corticomuscular coherence measure, respectively.

## Data Availability

The experimental datasets for this manuscript are not publicly available because raw data regarding human subjects (eg, genetic raw data, personal data) are not shared freely to protect the privacy of the human subjects involved in this study; no consent for open sharing has been obtained. Requests to access an anonymous data set and the simulated data should be directed to Christian Lassmann.

## Data Availability

The datasets for this manuscript are not publicly available because raw data regarding human subjects (e.g., genetic raw data, personal data) are not shared freely to protect the privacy of the human subjects involved in this study; no consent for open sharing has been obtained. Requests to access an anonymous data set and simulation data should be directed to Christian Lassmann.

## Authors Contribution

C.L. designed the work, acquired, analyzed, and interpreted the data, and wrote the manuscript.

W.I. interpreted the data, and wrote and revised the manuscript. T.W.R. interpreted the data and revised the manuscript. L.S. interpreted the data and revised the manuscript. M.G. interpreted the data and revised the manuscript. D.F.B.H. designed the work, interpreted the data, and wrote and revised the manuscript.

All authors approved the final version of the manuscript and agreed to be accountable for all aspects of the work in ensuring that questions related to the accuracy or integrity of any part of the work are appropriately investigated and resolved. All persons designated as authors qualify for authorship, and all those who qualify for authorship are listed.

## Acknowledgments

This work was supported by the Forum Ge(h)n mit HSP and the Förderverein für HSP-Forschung e.V. (grant to L.S. and T.W.R.). T.W.R. received funding from the University of Tübingen, medical faculty, for the Clinician Scientist Program Grant: #386–0-0. L.S. is a member of the European Reference Network for Rare Neurological Diseases, Project ID 739510.

## References

Bruel A, Ghorbel SB, Russo AD, Stanev D, Armand S, Courtine G, Ijspeert A. Investigation of neural and biomechanical impairments leading to pathological toe and heel gaits using neuromusculoskeletal modelling. The Journal of Physiology. 2022; n/a(n/a). doi: https://doi.org/10.1113/JP282609.

De Groote F, Falisse A. Perspective on musculoskeletal modelling and predictive simulations of human movement to assess the neuromechanics of gait. Proceedings of the Royal Society B: Biological Sciences. 2021; 288(1946):20202432. doi: https://doi.org/10.1098/rspb.2020.2432.

Delp SL, Anderson FC, Arnold AS, Loan P, Habib A, John CT, Guendelman E, Thelen DG. OpenSim: Open-Source Software to Create and Analyze Dynamic Simulations of Movement. IEEE Transactions on Biomedical Engineering. 2007; 54(11):1940–1950. doi: https://doi.org/10.1109/TBME.2007.901024.

Di Russo A, Stanev D, Armand S, Ijspeert A. Sensory modulation of gait characteristics in human locomotion: A neuromusculoskeletal modeling study. PLOS Computational Biology. 2021 05; 17(5):1–33. doi: 10.1371/journal.pcbi.1008594.

Fink JK. Hereditary spastic paraplegia. Current neurology and neuroscience reports. 2006; 6(1):65–76. doi: https://doi.org/10.1007/s11910-996-0011-1.

Fink JK. Hereditary spastic paraplegia: clinico-pathologic features and emerging molecular mechanisms. Acta neuropathologica. 2013; 126(3):307–328. doi: https://doi.org/10.1007/s00401-013-1115-8.

Geijtenbeek T. SCONE: Open Source Software for Predictive Simulation of Biological Motion. Journal of Open Source Software. 2019; 4(38):1421. doi: https://doi.org/10.21105/joss.01421.

Geijtenbeek T, The Hyfydy Simulation Software; 2021. https://hyfydy.com, https://hyfydy.com.

Geyer H, Herr H. A Muscle-Reflex Model That Encodes Principles of Legged Mechanics Produces Human Walking Dynamics and Muscle Activities. IEEE Transactions on Neural Systems and Rehabilitation Engineering. 2010; 18(3):263–273. doi: 10.1109/TNSRE.2010.2047592.

Globas C, du Montcel ST, Baliko L, Boesch S, Depondt C, DiDonato S, Durr A, Filla A, Klockgether T, Mariotti C, Melegh B, Rakowicz M, Ribai P, Rola R, Schmitz-Hubsch T, Szymanski S, Timmann D, Van de Warrenburg BP, Bauer P, Schols L. Early symptoms in spinocerebellar ataxia type 1, 2, 3, and 6. Movement Disorders. 2008; 23(15):2232–2238. doi: https://doi.org/10.1002/mds.22288.

Haeufle DFB, Schmortte B, Geyer H, Müller R, Schmitt S. The Benefit of Combining Neuronal Feedback and Feed-Forward Control for Robustness in Step Down Perturbations of Simulated Human Walking Depends on the Muscle Function. Frontiers in Computational Neuroscience. 2018; 12. doi: https://doi.org/10.3389/fncom.2018.00080.

Harding AE. CLASSIFICATION OF THE HEREDITARY ATAXIAS AND PARAPLEGIAS. The Lancet. 1983; 321(8334):1151–1155. doi: https://doi.org/10.1016/S0140-6736(83)92879-9, originally published as Volume 1, Issue 8334.

Hazan J, Fonknechten N, Mavel D, Paternotte C, Samson D, Artiguenave F, Davoine CS, Cruaud C, Dürr A, Wincker P, et al. Spastin, a new AAA protein, is altered in the most frequent form of autosomal dominant spastic paraplegia. Nature genetics. 1999; 23(3):296–303. doi: https://doi.org/10.1038/15472.

Igel C, Suttorp T, Hansen N. A Computational Efficient Covariance Matrix Update and a (1+1)-CMA for Evolution Strategies. In: Proceedings of the 8th Annual Conference on Genetic and Evolutionary Computation GECCO ‘06, New York, NY, USA: Association for Computing Machinery; 2006. p. 453–460. doi: https://doi.org/10.1145/1143997.1144082.

Ilg W, Fleszar Z, Schatton C, Hengel H, Harmuth F, Bauer P, Timmann D, Giese M, Schöls L, Synofzik M. Individual changes in preclinical spinocerebellar ataxia identified via increased motor complexity. Movement Disorders. 2016; 31(12):1891–1900. doi: https://doi.org/10.1002/mds.26835.

Jansen K, De Groote F, Aerts W, De Schutter J, Duysens J, Jonkers I. Altering length and velocity feedback during a neuro-musculoskeletal simulation of normal gait contributes to hemiparetic gait characteristics. Journal of neuroengineering and rehabilitation. 2014; 11(1):1–15. doi: https://doi.org/10.1186/1743-0003-11-78.

van der Krogt MM, Doorenbosch CA, Becher J, Harlaar J. Dynamic spasticity of plantar flexor muscles in cerebral palsy gait. Journal of rehabilitation medicine. 2010; 42(7):656–663. doi: https://doi.org/10.2340/16501977-0579.

van der Krogt MM, Bar-On L, Kindt T, Desloovere K, Harlaar J. Neuro-musculoskeletal simulation of instrumented contracture and spasticity assessment in children with cerebral palsy. Journal of NeuroEngineering and Rehabilitation. 2016; 13(1):1–11. doi: https://doi.org/10.1186/s12984-016-0170-5.

Lassmann C, Ilg W, Schneider M, Völker M, Haeufle DFB, Schüle R, Giese M, Synofzik M, Schöls L, Rattay TW. Specific gait changes in prodromal hereditary spastic paraplegia type 4 - preSPG4 study. Movement Disorders. 2022; doi: https://doi.org/10.1002/mds.29199.

Lindig T, Bender B, Hauser TK, Mang S, Schweikardt D, Klose U, Karle KN, Schüle R, Schöls L, Rattay TW. Gray and white matter alterations in hereditary spastic paraplegia type SPG4 and clinical correlations. Journal of neurology. 2015; 262(8):1961–1971. doi: https://doi.org/10.1007/s00415-015-7791-7.

Marsden J, Ramdharry G, Stevenson V, Thompson A. Muscle paresis and passive stiffness: Key determinants in limiting function in Hereditary and Sporadic Spastic Paraparesis. Gait & Posture. 2012; 35(2):266–271. doi: https://doi.org/10.1016/j.gaitpost.2011.09.018.

Martino G, Ivanenko Y, Serrao M, Ranavolo A, Draicchio F, Casali C, Lacquaniti F. Locomotor coordination in patients with Hereditary Spastic Paraplegia. Journal of Electromyography and Kinesiology. 2019; 45:61–69. doi: https://doi.org/10.1016/j.jelekin.2019.02.006.

Martino G, Ivanenko Y, Serrao M, Ranavolo A, Draicchio F, Rinaldi M, Casali C, Lacquaniti F. Differential changes in the spinal segmental locomotor output in Hereditary Spastic Paraplegia. Clinical Neurophysiology. 2018; 129(3):516–525. doi: https://doi.org/10.1016/j.clinph.2017.11.028.

Millard M, Uchida T, Seth A, Delp SL. Flexing Computational Muscle: Modeling and Simulation of Musculotendon Dynamics. Journal of Biomechanical Engineering. 2013 02; 135(2). doi: https://doi.org/10.1115/1.4023390, 021005.

Mirelman A, Bernad-Elazari H, Thaler A, Giladi-Yacobi E, Gurevich T, Gana-Weisz M, Saunders-Pullman R, Raymond D, Doan N, Bressman SB, Marder KS, Alcalay RN, Rao AK, Berg D, Brockmann K, Aasly J, Waro BJ, Tolosa E, Vilas D, Pont-Sunyer C, et al. Arm swing as a potential new prodromal marker of Parkinson’s disease. Movement Disorders. 2016; 31(10):1527–1534. doi: https://doi.org/10.1002/mds.26720.

Ong CF, Geijtenbeek T, Hicks JL, Delp SL. Predicting gait adaptations due to ankle plantarflexor muscle weakness and contracture using physics-based musculoskeletal simulations. PLOS Computational Biology. 2019 10; 15(10):1–27. doi: 10.1371/journal.pcbi.1006993.

Piccinini L, Cimolin V, D’Angelo MG, Turconi AC, Crivellini M, Galli M. 3D gait analysis in patients with hereditary spastic paraparesis and spastic diplegia: A kinematic, kinetic and EMG comparison. European Journal of Paediatric Neurology. 2011; 15(2):138–145. doi: https://doi.org/10.1016/j.ejpn.2010.07.009.

Rattay TW, Völker M, Rautenberg M, Kessler C, Wurster I, Winter N, Haack TB, Lindig T, Hengel H, Synofzik M, Schüle R, Martus P, Schöls L. The prodromal phase of hereditary spastic paraplegia type 4: the preSPG4 cohort study. Brain. 2022 04; doi: https://doi.org/10.1093/brain/awac155, awac155.

Rezende TJ, de Albuquerque M, Lamas GM, Martinez AR, Campos BM, Casseb RF, Silva CB, Branco LM, D’Abreu A, Lopes-Cendes I, et al. Multimodal MRI-based study in patients with SPG4 mutations. PLoS One. 2015; 10(2):e0117666. doi: https://doi.org/10.1371/journal.pone.0117666.

Rinaldi M, Ranavolo A, Conforto S, Martino G, Draicchio F, Conte C, Varrecchia T, Bini F, Casali C, Pierelli F, Serrao M. Increased lower limb muscle coactivation reduces gait performance and increases metabolic cost in patients with hereditary spastic paraparesis. Clinical Biomechanics. 2017; 48:63–72. doi: https://doi.org/10.1016/j.clinbiomech.2017.07.013.

Schreff L, Haeufle DF, Vielemeyer J, Müller R. Evaluating anticipatory control strategies for their capability to cope with step-down perturbations in computer simulations of human walking. Scientific reports. 2022; 12(1):1–11. doi: https://doi.org/10.1038/s41598-022-14040-0.

Schüle R, Holland-Letz T, Klimpe S, Kassubek J, Klopstock T, Mall V, Otto S, Winner B, Schöls L. The Spastic Paraplegia Rating Scale (SPRS). Neurology. 2006; 67(3):430–434. doi: 10.1212/01.wnl.0000228242.53336.90.

Serrao M, Rinaldi M, Ranavolo A, Lacquaniti F, Martino G, Leonardi L, Conte C, Varrecchia T, Draicchio F, Coppola G, Casali C, Pierelli F. Gait Patterns in Patients with Hereditary Spastic Paraparesis. PLOS ONE. 2016 10; 11(10):1–16. doi: 10.1371/journal.pone.0164623.

Song S, Geyer H. A neural circuitry that emphasizes spinal feedback generates diverse behaviours of human locomotion. The Journal of physiology. 2015; 593(16):3493–3511. doi: https://doi.org/10.1113/JP270228.

Van Lith BJ, den Boer J, van de Warrenburg BP, Weerdesteyn V, Geurts A. Functional effects of botulinum toxin type A in the hip adductors and subsequent stretching in patients with hereditary spastic paraplegia. Journal of Rehabilitation Medicine. 2019; 51. doi: https://doi.org/10.2340/16501977-2556.

Veerkamp K, Waterval NFJ, Geijtenbeek T, Carty CP, Lloyd DG, Harlaar J, van der Krogt MM. Evaluating cost function criteria in predicting healthy gait. Journal of Biomechanics. 2021; 123:110530. doi: https://doi.org/10.1016/j.jbiomech.2021.110530.

Wang JM, Hamner SR, Delp SL, Koltun V. Optimizing Locomotion Controllers Using Biologically-Based Actuators and Objectives. ACM Trans Graph. 2012 jul; 31(4). doi: https://doi.org/10.1145/2185520.2185521.

Waterval NFJ, Veerkamp K, Geijtenbeek T, Harlaar J, Nollet F, Brehm MA, van der Krogt MM. Validation of forward simulations to predict the effects of bilateral plantarflexor weakness on gait. Gait & Posture. 2021; 87:33–42. doi: https://doi.org/10.1016/j.gaitpost.2021.04.020.

