## Supplementary material for "Dysfunctional neuro-muscular mechanisms explain gradual gait changes in prodromal spastic paraplegia" (from another version of this preprint: version 1, 2022-10-17)

**Christian Laßmann**[1,2,3*,§], **Winfried Ilg**[2,4], **Tim W. Rattay**[5,6,7], **Ludger Schöls**[5,6,7], **Martin Giese**[2,4], **Daniel F.B. Haeufle**[1,4,8]

***For correspondence:**
 (FMS)

**Present address:** §
Multi-level Modeling in Motor Control and Rehabilitation Robotics
Hertie Institute for Clinical Brain Research
Otfried-Müller-Str. 25
72076 Tübingen, Germany


[1]Multi-level Modeling in Motor Control and Rehabilitation Robotics, Hertie Institute for Clinical Brain Research, University of Tübingen, Tübingen, Germany; [2]Section Computational Sensomotorics, Hertie Institute for Clinical Brain Research, Tübingen, Germany; [3]Department of Computer Engineering, Wilhelm-Schickard-Institute for Computer Science, University of Tübingen, Tübingen, Germany; [4]Centre for Integrative Neuroscience (CIN), Tübingen, Germany; [5]Department of Neurodegenerative Disease, Hertie-Institute for Clinical Brain Research, and Center for Neurology, University of Tübingen, Tübingen, Germany; [6]German Center for Neurodegenerative Diseases (DZNE), Tübingen, Germany; [7]Center for rare diseases (ZSE) University of Tübingen, Tübingen, Germany; [8]Institute for Modeling and Simulation of Biomechanical Systems, University of Stuttgart, Stuttgart, Germany

### Appendix

#### Velocity feedback gain and muscle weakness parameters

**Corresponding values of muscle weakness and velocity feedback gain for simulation iterations $\omega$.**

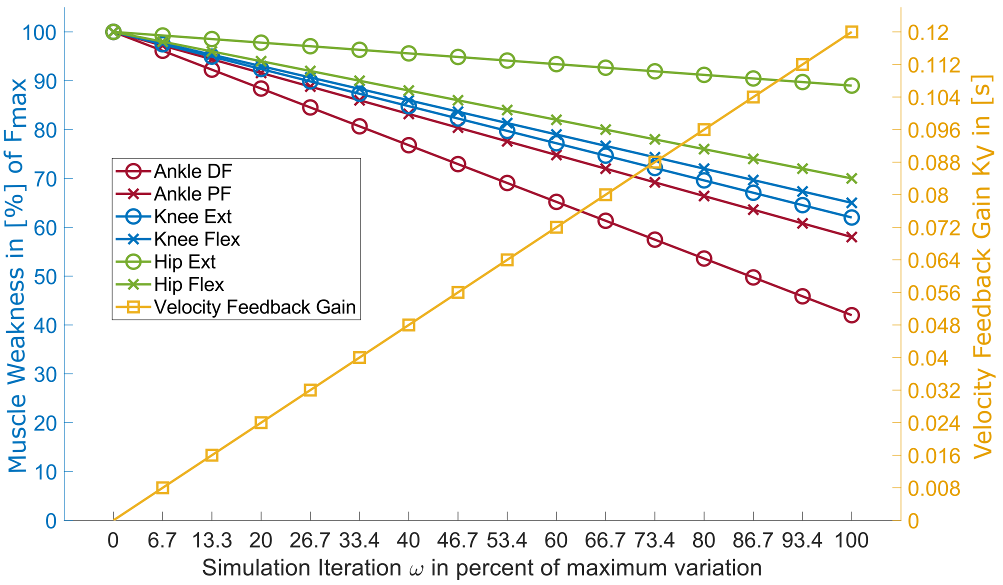


**Appendix 0 Figure 1.** Velocity feedback gains for SOL, GAS and TA and percentage of maximum isometric forces for plantarflexors, dorsiflexors, knee, and hip flexors and extensors, with their relative change in $\omega$. DF ≡ Dorsiflexion, PF ≡ Plantarflexion, Ext ≡ Extension, and Flex ≡ Flexion.

#### Increasing velocity feedback gain

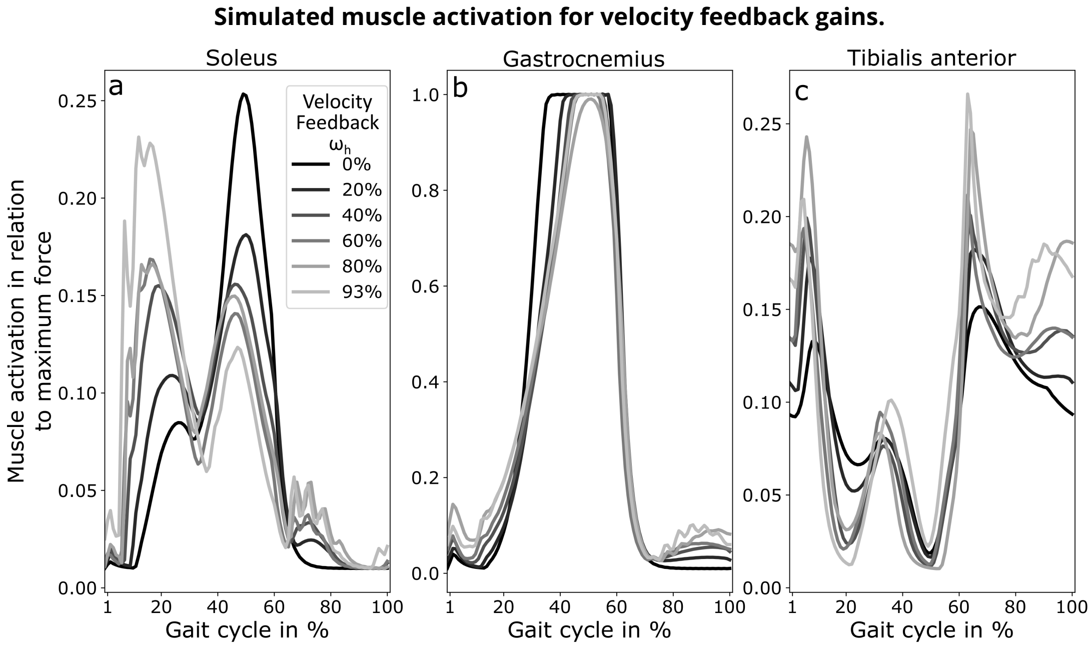


**Appendix 0 Figure 2.** Muscle activation of Soleus, Gastrocnemius and Tibialis anterior over the gait cycle in percent for different levels of velocity feedback gain.

**Kinematic features for velocity feedback gain results over all iterations and correlation.**

| **Velocity Feedback $\omega_h$** | Gait Speed | Stride Length | ankle RoM | Min PF | ankle at HS | ankle at max HGC | knee RoM | knee max angle | knee at HS |
|---|---|---|---|---|---|---|---|---|---|
| 0% | 1.20 | 1.48 | 25.43 | -17.30 | 7.24 | -14.70 | 60.99 | 62.27 | 3.59 |
| 6.7% | 1.05 | 1.31 | 19.74 | -11.67 | 3.99 | -9.66 | 65.00 | 66.42 | 3.30 |
| 13.3% | 1.10 | 1.36 | 19.60 | -12.67 | 3.83 | -8.95 | 63.13 | 64.98 | 3.50 |
| 20% | 1.16 | 1.44 | 19.00 | -11.45 | 2.31 | -8.05 | 61.69 | 63.02 | 3.37 |
| 26.7% | 1.07 | 1.35 | 16.25 | -8.70 | 0.95 | -5.75 | 62.29 | 63.34 | 3.11 |
| 33.3% | 1.10 | 1.43 | 16.99 | -9.70 | -0.02 | -7.38 | 62.72 | 62.88 | 4.17 |
| 40% | 1.05 | 1.35 | 16.22 | -8.35 | -0.51 | -5.11 | 61.92 | 63.36 | 3.79 |
| 46.7% | 1.03 | 1.33 | 15.77 | -7.81 | -1.35 | -3.93 | 62.94 | 64.12 | 3.82 |
| 53.3% | 1.05 | 1.34 | 15.93 | -4.29 | -0.74 | -2.24 | 64.35 | 65.51 | 9.73 |
| 60% | 1.08 | 1.39 | 15.69 | -6.17 | -1.10 | -3.27 | 62.22 | 62.70 | 11.50 |
| 66.7% | 1.08 | 1.39 | 15.30 | -7.78 | -2.29 | -3.97 | 62.79 | 63.44 | 9.57 |
| 73.3% | 1.04 | 1.34 | 14.86 | -5.82 | -1.65 | -1.98 | 60.87 | 63.10 | 12.66 |
| 80% | 1.03 | 1.32 | 15.09 | -7.35 | -4.42 | -3.52 | 62.97 | 63.95 | 4.26 |
| 86.7% | 1.05 | 1.38 | 14.12 | -5.04 | -1.67 | -2.32 | 61.03 | 61.73 | 13.01 |
| 93% | 1.06 | 1.33 | 13.72 | -4.78 | -0.73 | -2.32 | 65.52 | 67.12 | 18.35 |
| **Correlation**<br>Iteration 0 - 14 | rho=-0.53<br>p=0.04* | rho=-0.32<br>p=0.25 | rho=-0.99<br>p<0.001*** | rho=0.9<br>p<0.001*** | rho=-0.87<br>p<0.001*** | rho=0.88<br>p<0.001*** | rho=0.06<br>p=0.82 | rho=0.06<br>p=0.84 | rho=0.88<br>p=0.001*** |

**Appendix 0 Table 1.** Shown are all walking simulations for gradual increasing velocity feedback gains. In the bottom row pearson correlation coefficients for the respective features are shown. RoM ≡ Range of Motion, PF ≡ Plantarflexion, HS ≡ Heel strike, and HGC ≡ Heel ground clearance.

#### Increasing muscle weakness

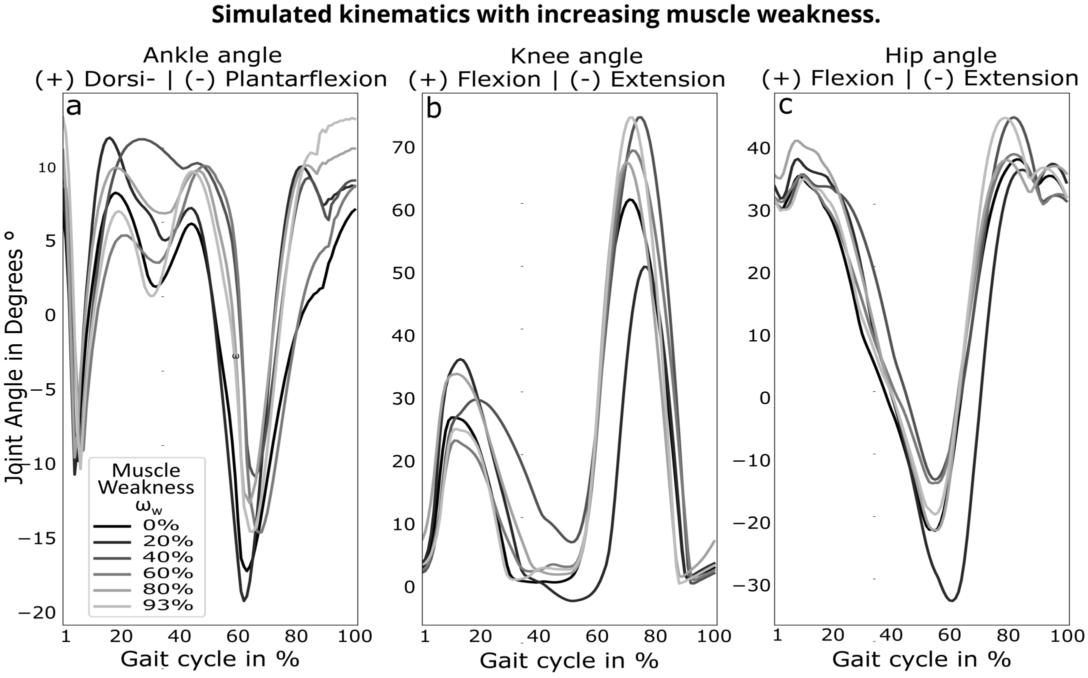


**Appendix 0 Figure 3.** Flexion and extension angles of the ankle, knee, and hip joints over the gait cycle in percent for different levels of muscle weakness. For more details on different gait features see ***Table 2***

**Kinematic features for muscle weakness results over all iterations and correlation.**

| **Weakness** $\omega_w$ | Gait Speed | Stride Length | ankle RoM | Min PF | ankle at HS | ankle at max HGC | knee RoM | knee max angle | knee at HS |
|---|---|---|---|---|---|---|---|---|---|
| 0% | 1.20 | 1.48 | 25.43 | -17.30 | 7.24 | -14.70 | 60.99 | 62.27 | 3.59 |
| 6.7% | 1.18 | 1.45 | 26.80 | -15.50 | 8.58 | -12.55 | 66.13 | 67.85 | 3.65 |
| 13.3% | 1.10 | 1.35 | 24.89 | -14.98 | 9.30 | -10.24 | 68.55 | 69.89 | 3.54 |
| 20% | 1.07 | 1.54 | 31.11 | -19.05 | 8.83 | -4.98 | 53.30 | 52.06 | 4.30 |
| 26.7% | 0.96 | 1.18 | 21.00 | -12.71 | 4.89 | -9.04 | 72.13 | 73.37 | 3.42 |
| 33.3% | 1.13 | 1.42 | 26.78 | -17.66 | 8.32 | -14.76 | 66.48 | 67.88 | 3.56 |
| 40% | 1.14 | 1.33 | 22.84 | -10.82 | 9.24 | -3.64 | 74.37 | 75.43 | 2.77 |
| 46.7% | 0.98 | 1.18 | 22.44 | -12.97 | 8.49 | -9.73 | 73.02 | 74.22 | 2.96 |
| 53.3% | 0.96 | 1.45 | 28.33 | -9.50 | 11.66 | 2.46 | 61.01 | 61.93 | 3.81 |
| 60% | 1.10 | 1.33 | 24.72 | -14.52 | 8.92 | -12.91 | 68.46 | 70.41 | 3.27 |
| 66.7% | 1.08 | 1.32 | 24.38 | -13.80 | 10.63 | -10.50 | 74.27 | 75.48 | 3.63 |
| 73.3% | 1.07 | 1.28 | 25.85 | -14.87 | 10.63 | -12.78 | 72.90 | 74.42 | 3.14 |
| 80% | 1.14 | 1.50 | 23.94 | -12.59 | 11.34 | -9.08 | 66.08 | 68.18 | 7.95 |
| 86.7% | 0.99 | 1.30 | 26.83 | -15.66 | 11.19 | -12.50 | 68.20 | 69.68 | 3.58 |
| 93.3% | 1.11 | 1.42 | 27.84 | -14.48 | 13.42 | -9.36 | 74.30 | 75.43 | 3.97 |
| 100% | 1.11 | 1.51 | 28.65 | -14.63 | 12.16 | -11.72 | 65.65 | 67.87 | 7.63 |
| **Correlation** Iteration 0 - 15 | rho=-0.19 p=0.47 | rho=-0.09 p=0.74 | rho=0.19 p=0.48 | rho=0.33 p= 0.213 | rho=0.8 p<0.001*** | rho=0.06 p=0.81 | rho= 0.25 p=0.35 | rho=0.36 p=0.18 | rho=0.26 p=0.33 |

**Appendix 0 Table 2.** Shown are all walking simulations for gradual increasing muscle weakness. In the bottom row pearson correlation coefficients for the respective features are shown. RoM ≡ Range of Motion, PF ≡ Plantarflexion, HS ≡ Heel strike, and HGC ≡ Heel ground clearance.

#### Increasing velocity feedback gain and muscle weakness

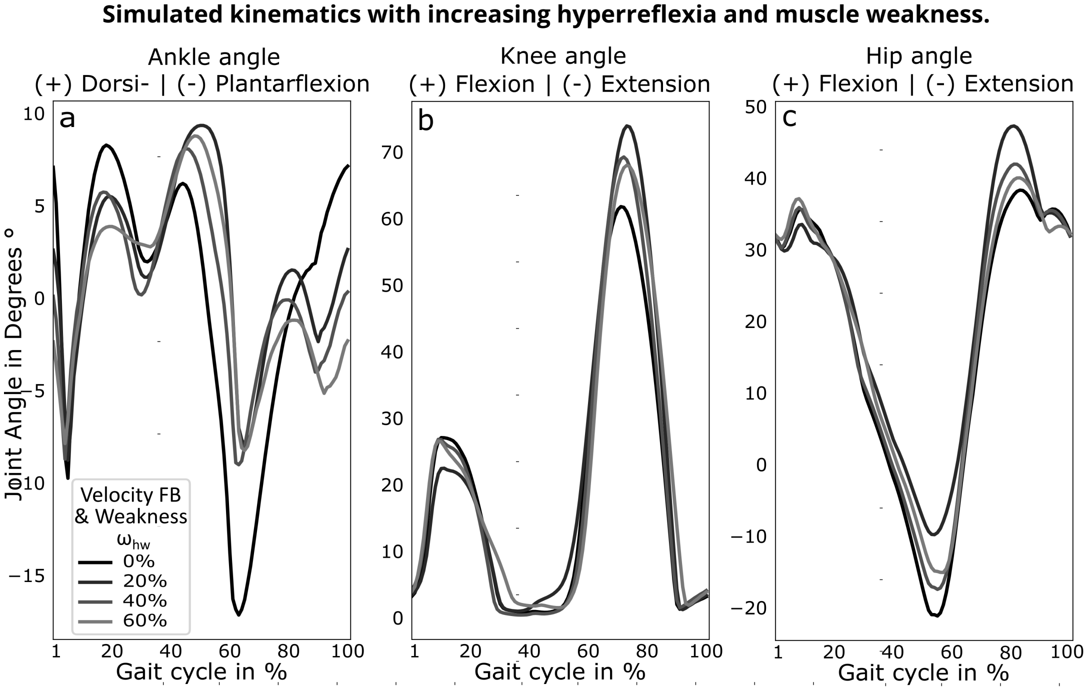


**Appendix 0 Figure 4.** Flexion and extension angles of the ankle, knee, and hip joints over the gait cycle in percent for velocity feedback gain and muscle weakness. For more details on different gait features see ***Table 3***

**Kinematic features for velocity feedback gain and muscle weakness results over all iterations and correlation.**

| **Velocity FB + Weakness $\omega_{hw}$** | Gait Speed | Stride Length | ankle RoM | Min PF | ankle at HS | ankle at max HGC | knee RoM | knee max angle | knee at HS |
|---|---|---|---|---|---|---|---|---|---|
| 0% | 1.20 | 1.48 | 25.43 | -17.30 | 7.24 | -14.70 | 60.99 | 62.27 | 3.59 |
| 6.7% | 1.21 | 1.40 | 20.03 | -13.32 | 1.59 | -8.47 | 64.36 | 66.82 | 2.45 |
| 13.3% | 1.06 | 1.33 | 21.45 | -13.37 | 2.87 | -10.72 | 63.23 | 66.52 | 3.50 |
| 20% | 0.96 | 1.20 | 18.23 | -8.67 | 2.56 | -2.98 | 72.93 | 74.56 | 3.74 |
| 26.7% | 1.12 | 1.36 | 18.60 | -9.64 | 0.04 | -5.96 | 65.40 | 67.29 | 3.40 |
| 33.3% | 1.05 | 1.42 | 17.61 | -9.27 | 3.45 | -5.44 | 62.23 | 63.39 | 5.80 |
| 40% | 1.03 | 1.33 | 17.26 | -9.52 | 0.25 | -3.89 | 68.82 | 69.94 | 4.55 |
| 46.7% | 1.07 | 1.40 | 17.38 | -8.30 | 0.10 | -5.45 | 64.83 | 65.70 | 5.68 |
| 53.3% | 1.06 | 1.30 | 16.74 | -7.22 | 2.09 | -1.84 | 73.45 | 74.70 | 4.34 |
| 60% | 0.98 | 1.26 | 17.11 | -8.40 | -2.21 | -4.88 | 66.54 | 69.26 | 4.33 |
| 66.7% | | | | | | | | | |
| 73.3% | 0.96 | 1.46 | 56.11 | -70.33 | -36.15 | -31.53 | 72.69 | 66.19 | 2.06 |
| **Correlation** Iteration 0 - 9 | rho=-0.52 p=0.13 | rho=-0.26 p=0.47 | rho=-0.98 p<0.001*** | rho=0.96 p<0.001*** | rho=-0.96 p<0.001*** | rho=0.98 p<0.001*** | rho=0.12 p=0.75 | rho=0.01 p=0.99 | rho=0.76 p=0.011* |

**Appendix 0 Table 3.** Shown are all walking simulations for gradual increasing velocity feedback gains and muscle weakness. In the bottom row pearson correlation coefficients for the respective features are shown. RoM ≡ Range of Motion, PF ≡ Plantarflexion, HS ≡ Heel strike, and HGC ≡ Heel ground clearance.

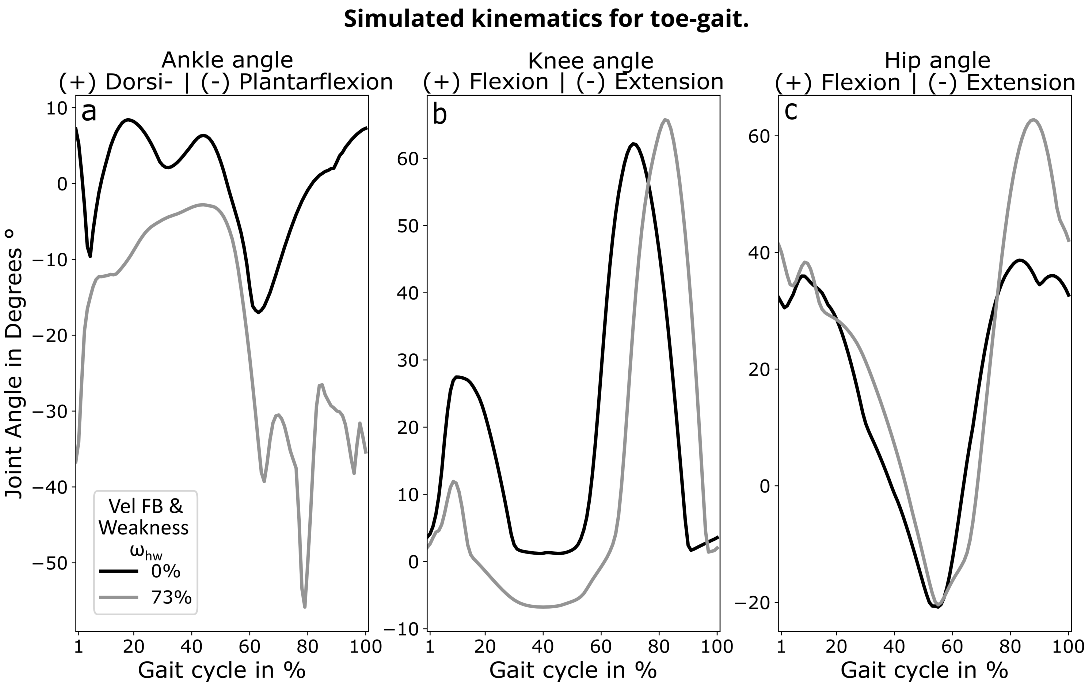


**Appendix 0 Figure 5.** Flexion and extension angles of ankle, knee, and hip joints over the gait cycle in percent for velocity feedback gain and muscle weakness of $\omega_{hw} = 73\%$ and $\omega_{hw} = 0\%$ for comparison. For this scenario, the optimization process produced a controller with stable toe gait with initial ball contact. For more details on different gait features see ***Table 3***

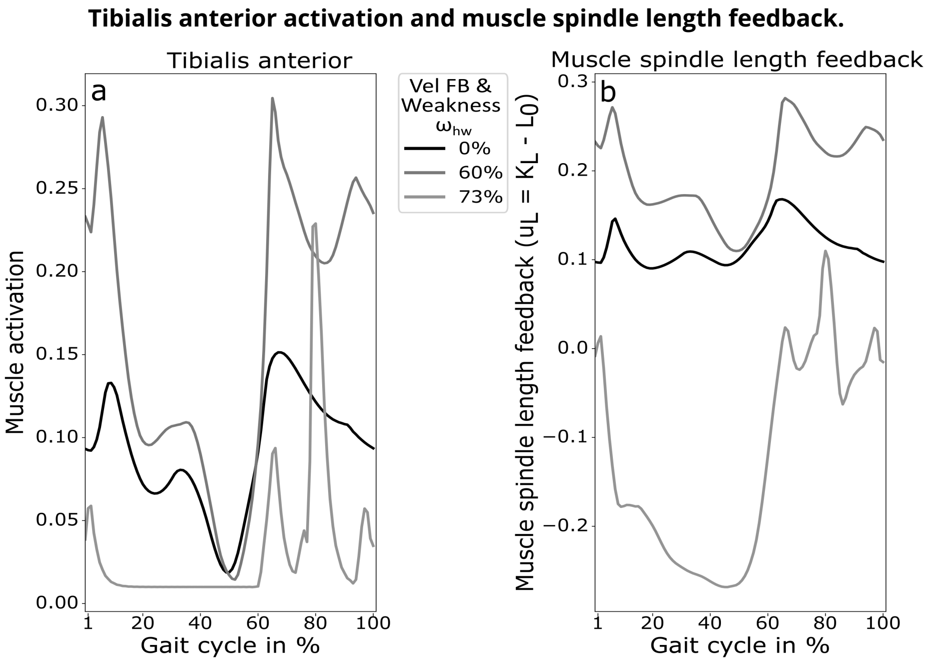


**Appendix 0 Figure 6.** TA activation and muscle spindle length feedback over the gait cycle in percent for different levels of combined velocity feedback and muscle weakness ($\omega_{hw} = 0\%$, no alteration; $\omega_{hw} = 60\%$ heel strike gait; and $\omega_{hw} = 73\%$ toe-gait). For $\omega_{hw} = 73\%$ the muscle spindle length offset ($L_0$) is increased in comparison to other scenarios, which reduces TA activation and reciprocal inhibition of plantarflexor muscles.

#### Optimized parameters

**Correlation of optimization parameters for each simulation experiment.**

| **Condition** | Optimization | Effort | TA[$K_L$] | TA[$L_0$] | TA-SOL[$K_F$] | SOL[$K_F$] | GAS[$K_F$] | VAS[$K_F$] | VAS[$C_0$] |
|---|---|---|---|---|---|---|---|---|---|
| VelFB | rho=0.78, p=0.001*** | rho=0.79, p<0.001*** | rho=0.72, p=0.002** | rho=0.49, p=0.064 | rho=-0.03, p=0.919 | rho=-0.94, p<0.001*** | rho=-0.94, p<0.001*** | rho=-0.44, p=0.098 | rho=-0.5, p=0.056 |
| Weakness | rho=-0.02, p=0.931 | rho=-0.02, p=0.931 | rho=0.41, p=0.113 | rho=-0.01, p=0.957 | rho=0.27, p=0.311 | rho=0.54, p=0.031* | rho=-0.23, p=0.399 | rho=0.34, p=0.2 | rho=0.0, p=1.0 |
| VelFB + Weakness | rho=0.63, p=0.05 | rho=0.66, p=0.038* | rho=0.47, p=0.174 | rho=0.39, p=0.26 | rho=0.48, p=0.162 | rho=-0.98, p<0.001*** | rho=-0.99, p<0.001*** | rho=-0.48, p=0.162 | rho=-0.84, p=0.002** |

**Appendix 0 Table 4.** Shown are all walking simulations for gradual increasing velocity feedback gains and muscle weakness. In the bottom row pearson correlation coefficients for the respective features are shown. $K_L \equiv$ muscle spindle length feedback gain, $L_0 \equiv$ length feedback offset, $K_F \equiv$ muscle spindle force feedback gain, and $C_0 \equiv$ constant muscle actuation.
